# Multi-expert consensus annotations of spontaneous and stimulation-induced seizures in stereotactic EEG

**DOI:** 10.64898/2026.01.15.26344025

**Authors:** William K.S. Ojemann, Daniel J. Zhou, Catherine V. Kulick-Soper, Jacob Korzun, Joshua J. LaRocque, Caren Armstrong, Benjamin C. Kennedy, Sudha K. Kessler, Eric Marsh, Nishant Sinha, Brian Litt, Erin Conrad

## Abstract

Automated seizure detection and localization from intracranial EEG requires validated benchmark datasets with expert annotations, yet existing open datasets lack multi-expert consensus annotations and exclude stimulation-induced seizures. We present stereotactic EEG recordings from 83 seizures (46 spontaneous, 37 stimulation-induced) across 32 patients (19 from the University of Pennsylvania, 13 from the Children’s Hospital of Philadelphia) with drug-resistant epilepsy. Three board-certified epileptologists independently annotated each seizure for onset time, onset channels, and channels seizing at 10 seconds post-onset using a standardized protocol. Consensus annotations were determined through majority voting. Inter-rater agreement was κ = 0.64 for onset channels and κ = 0.62 for spread channels. Individual rater agreement with consensus was κ = 0.81 for onset and κ = 0.80 for spread. Agreement metrics did not differ between spontaneous and stimulation-induced seizures. All data follow Brain Imaging Data Structure (BIDS) standards and include electrode localizations, patient demographics, and clinical outcomes. This dataset enables the validation of seizure onset and spread detection and localization against human expert performance and supports comparative analysis of seizure networks across spontaneous and stimulation-induced seizures.

## BACKGROUND & SUMMARY

Intracranial EEG (iEEG) remains the gold standard for localizing the seizure onset zone (SOZ) in patients with drug-resistant focal epilepsy being evaluated for surgery.^1,2^ Identifying the SOZ and early spread regions through iEEG monitoring guides surgical planning, including resection boundaries, sites for laser ablation, or placement of neuromodulation devices. Currently, seizures are identified and annotated through visual review by epileptologists, who mark both the timing of seizure onset and the spatial distribution of ictal activity across implanted electrode contacts. This manual annotation process is labor-intensive, requiring expert attention to recordings of hundreds of channels at high sampling rates over continuous monitoring periods typically spanning days to weeks.^2^

Given the labor demands and potential variability of manual review, automated methods for detecting and localizing seizure onset and spread have become an active research priority. Machine learning approaches, ranging from spectral feature analysis and high frequency oscillation detection to deep learning architectures, show promise for augmenting or replacing manual annotation.^3–5^ However, the development and validation of these computational methods are fundamentally limited by the lack of openly available iEEG datasets with expert-validated ground truth annotations. Existing algorithms are typically trained and tested on private institutional datasets or validated against single clinician annotations, making cross-algorithm comparison difficult and reproducibility limited.^6^ Without standardized benchmark datasets, the field cannot establish baseline performance metrics or systematically evaluate which approaches generalize across patients, institutions, and seizure types.

The quality of algorithmic training data is further complicated by the fact that manual annotation reliability itself is poorly characterized. One systematic review found only one study examining interrater reliability of SOZ identification from iEEG, reporting 97% agreement and Cohen’s kappa of 0.80 in nine patients with two reviewers.^7,8^ An additional study found poor intra- and interrater reliability regardless of sampling rate.^9^ A a recent study of 20 spontaneous seizures in 10 patients found moderate agreement for high-density (κ=0.49) and low-density (κ=0.45) iEEG recordings across six expert reviewers.^10^ Individual annotators may also vary in their interpretation and description of ambiguous seizure onset patterns, which also lacks standardization across clinical practice and research publications.^11^ Single-rater annotations, while clinically necessary, may therefore be insufficient as algorithmic ground truth. Algorithms trained on single annotations risk learning annotator-specific interpretation patterns rather than generalizable features of seizure activity. Multi-rater consensus annotations address this by identifying regions of expert agreement and quantifying uncertainty in ambiguous cases, while also establishing realistic performance benchmarks.

Several public iEEG datasets exist, including those from the UPenn and Mayo Clinic Seizure Detection Challenge, which have advanced automated seizure detection research.^12–14^ However, these datasets focus primarily on binary seizure detection (seizure present vs. absent) rather than spatial localization of onset and spread. Additionally, most available datasets contain only spontaneous seizures, excluding seizures induced by direct cortical electrical stimulation. Stimulation-induced (stim) seizures are increasingly being recognized as potentially valuable for refining SOZ localization and predicting surgical outcomes.^15,16^ To our knowledge, no existing open dataset provides multi-expert consensus annotations of both seizure onset channels and time-resolved spread patterns, nor do they include sufficient stim seizures to enable comparative analysis. This gap limits development of algorithms that can localize epileptogenic regions across different seizure triggers and characterize how ictal networks evolve over different temporal phases.

Here, we present the first open dataset of stereotactic EEG (sEEG) recordings with gold-standard consensus annotations of both seizure onset zones and 10-second spread patterns based on 83 seizures (46 spontaneous, 37 stimulation-induced) from 32 patients at the University of Pennsylvania (UPenn) and Children’s Hospital of Philadelphia (CHOP). Each seizure was independently annotated by three board-certified epileptologists following a standardized protocol, with consensus determined through majority voting. All data are formatted according to the Brain Imaging Data Structure (BIDS) standard and made publicly available via the Pennsieve platform.^17^

These data were compiled as part of our recent study on developing unsupervised seizure annotation tools and a quantitative comparison of stimulation-induced and spontaneous seizures.^18^ By publishing this dataset, we address multiple critical needs in computational epilepsy research. First, it provides a benchmark for validating automated seizure localization algorithms against multi-expert consensus rather than single annotations, enabling direct comparison of algorithm performance to human interrater reliability. Second, the inclusion of both spontaneous and stim seizures enables systematic comparison of localization patterns and network dynamics across seizure types, addressing questions about whether algorithms trained on spontaneous seizures generalize to stim events. Third, the dual annotations (onset and 10-second spread) allow development and testing of models that capture seizure propagation patterns, providing insight into the focality of epileptic networks. The included inter-rater reliability statistics establish baseline human performance levels for sEEG onset and propagation patterns for both spontaneous and stim seizures, providing realistic benchmarks and identifying characteristics associated with annotation uncertainty. Beyond computational applications, this represents the first systematic quantification of inter-expert agreement for spatial seizure localization, with implications for clinical interpretation and surgical decision-making.

## METHODS

### Participants

We included data from 32 patients (19 from the Hospital of the University of Pennsylvania and 13 from the Children’s Hospital of Philadelphia) who were implanted with sEEG depth electrodes as part of their presurgical evaluation for drug-resistant epilepsy between 2021 and 2024. The study was approved by the Institutional Review Boards at the Hospital of the University of Pennsylvania (HUP) and the Children’s Hospital of Philadelphia (CHOP). All participants provided written informed consent.

Patient demographics and clinical characteristics are summarized in **Table 1**. The cohort included 18 females (56%) and 14 males (44%). Age at epilepsy onset had a median of 14 years (interquartile range [IQR] 8-22). Epilepsy duration at the time of implantation had a median of 9 years (IQR 2.8-13.4). Clinical evaluation classified 20 patients (63%) as having unifocal seizure onset, 19 patients (59%) as having mesial temporal lobe epilepsy (MTLE), and 15 patients (47%) as having lesional epilepsy based on presurgical imaging. Surgical outcomes were available for 15 patients (47%), with 6 patients (40%) achieving Engel class I (seizure-free) outcomes. Detailed individual patient characteristics are provided in the participants.tsv file of our dataset.

**Table 1.**

| Cohort | Value |
| --- | --- |
| Total patients | 32 |
| University of Pennsylvania, n (%) | 19 (59%) |
| Children's Hospital of Philadelphia, n (%) | 13 (41%) |
| Demographics |  |
| Female, n (%) | 18 (56%) |
| Male, n (%) | 14 (44%) |
| Age at epilepsy onset, median (IQR) | 14 (8-22) |
| Epilepsy duration, median (IQR) | 9 (2.8-13.4) |
| Clinical characteristics |  |
| Unifocal seizure onset, n (%) | 20 (63%) |
| Mesial temporal lobe epilepsy, n (%) | 19 (59%) |
| Lesion on MRI, n (%) | 15 (47%) |
| Engel 1 outcome, n (%)* | 11 (40%) |
| Seizure data |  |
| Total seizures | 83 |
| Spontaneous seizures, n (%) | 46 (55%) |
| Stimulation-induced seizures, n (%) | 37 (45%) |

A total of 83 seizures were included in the dataset: 46 spontaneous seizures (55%) and 37 stim seizures (45%). All seizures were independently annotated by three or more board-certified expert epileptologists following a standardized protocol. Consensus annotations were determined through majority voting, as described in Technical Validation.

### iEEG Recordings

For each patient, subdural grid, strip, and/or depth electrodes were surgically implanted in different brain regions (HUP: manufactured by Ad-Tech corporation, Oak Creek, WI; CHOP: manufactured by PMT corporation, MN, USA or DIXI medical, USA), which were clinically selected based on case-specific presurgical evaluation. Signals were recorded using a Natus Quantum system (sampling rate: 512–2048 Hz) and referenced to an electrode contact hypothesized to be in non-epileptogenic tissue, typically medullary bone. The patient was then evaluated in the Epilepsy Monitoring Unit (EMU), where the electrodes were connected to an amplifier by an EEG technician, and the EEG signals were recorded and stored in a centralized database (**Fig. 1**).

**Figure 1:**
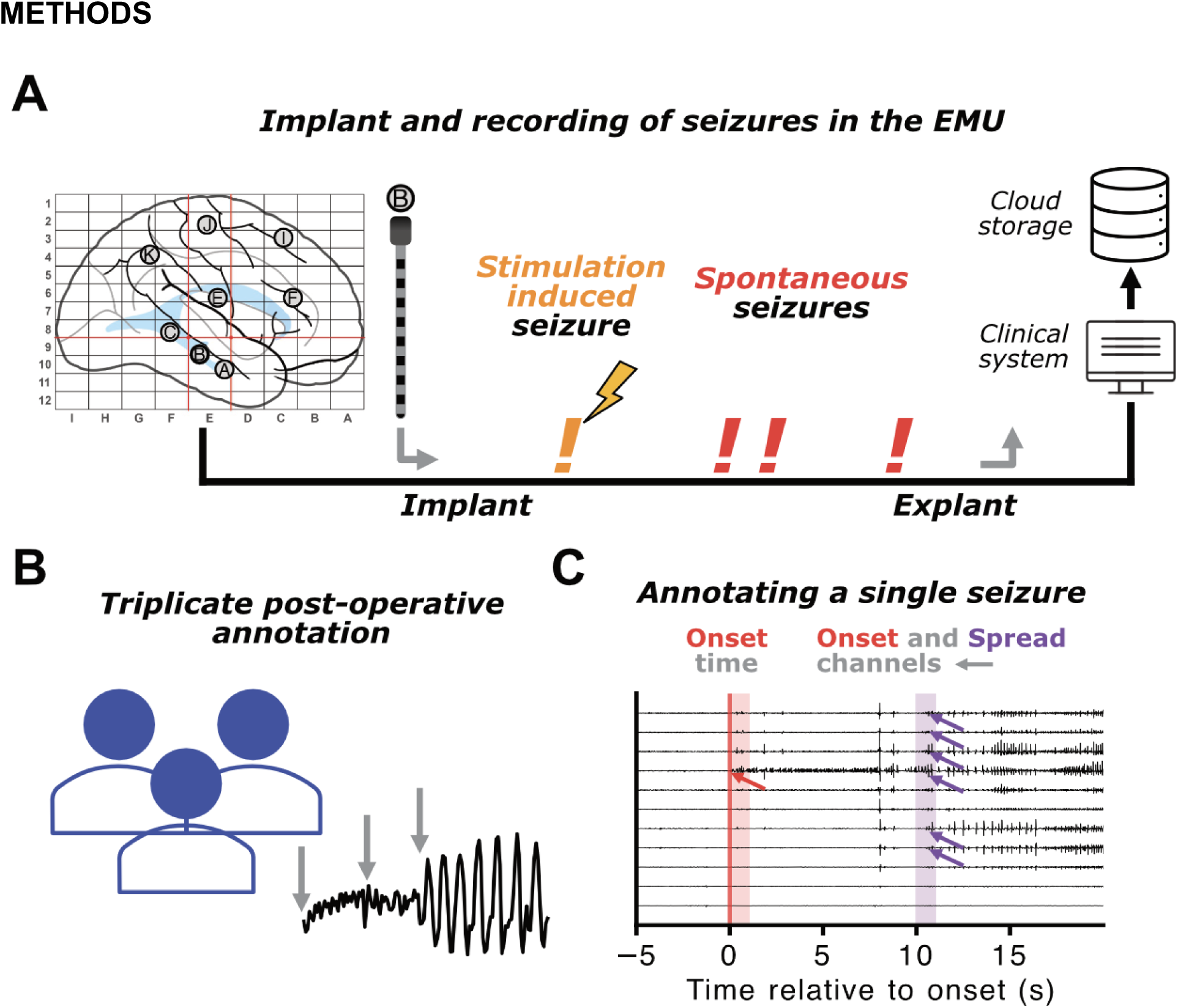
Intracranial EEG recording and annotation of seizure activity. **A)** Pipeline for EEG acquisition. Patients are implanted with depth electrodes to record EEG activity during a multi-day evaluation phase in the epilepsy monitoring unit (EMU). Shortly after implantation, patients undergo low-frequency stimulation to induce seizures, and are continuously monitored for spontaneously occurring seizures as anti-seizure medications are withdrawn. After sufficient electrographic information is recorded to make a clinical decision, electrodes are explanted, and continuously recorded EEG is uploaded to a cloud sharing platform. **B)** Three independent expert epileptologists annotated seizure onset time and EEG channels. **C)** The epileptologists were instructed to annotate seizure onset time (red bar), and involved channels seizing within 1 second from onset (red overlay and arrows), as well as channels seizing ten seconds after onset (purple overlay and arrows) during the seizure “spread” phase.

HUP patients underwent bipolar, biphasic stimulation at 1 Hz frequency, 3 mA amplitude, and pulse widths of 300 μs (initial 14 patients) or 500 μs (subsequent patients), yielding charge densities of 11.1-18.5 μC/cm². CHOP patients received stimulation with variable parameters: amplitudes of 1-8 mA, frequencies of 1-2 Hz, pulse widths of 300-500 μs, and charge densities of 17.9-59.69 μC/cm². The stimulation protocol targeted all adjacent bipolar contact pairs positioned in neural tissue that did not produce significant EEG artifacts. Time constraints occasionally necessitated stimulating alternating adjacent contact pairs (50% of total pairs). Stimulation was typically discontinued following the induction of an impaired awareness seizure.

### Electrode Localization

For patients from CHOP, the post-implant head CT was co-registered to the pre-implant brain MRI using Gardel software^19^ with subsequent atlas segmentation to the Desikan-Killiany (DK) atlas performed in Freesurfer.^20^ For HUP patients, we applied the iEEG-recon software^21^ to localize iEEG contacts in pre-implant T1 MRI space and MNI 152 space. Briefly, we segmented the pre-implant T1 MRI using FreeSurfer, applied the Desikan–Killiany atlas parcellation, co-registered the post-implant CT to the pre-implant T1 MRI, and localized electrode contacts in the native pre-implant T1 MRI space as well as in MNI space. All outputs and registrations were quality-checked for accuracy.

### Manual Annotations

Clinically identified electrographic and electroclinical seizures were manually annotated on ieeg.org by multiple experts trained in reading intracranial EEGs.^22^ These included spontaneous seizures, which occur without a clearly identifiable trigger, and stim seizures, which were elicited by low-frequency (1 Hz) electrical stimulation to certain leads in the brain. Each expert was instructed to follow a standardized EEG reviewing and annotation protocol.

Experts reviewed each EEG in bipolar longitudinal montage with all channels shown but with the option to hide any selected channels. For stim seizures, if the DC drift and stimulation artifact made the EEG unreadable, experts optionally applied a 3-50 Hz bandpass filter. If there is too much 60 Hz artifact, experts optionally applied a 59-61 Hz bandpass (notch) filter.

For each spontaneous or stim seizure, the expert marked the time of seizure onset as the unequivocal electrographic onset (UEO). The UEO represents the first time point during which there is a clear beginning to a continuous ictal pattern on EEG.^23^ If there are electrographic changes that could suggest a seizure onset but the pattern does not remain continuous with the rest of the seizure, then the expert was instructed not to mark the time point as UEO. For example, if there appeared to be a herald spike in certain leads, followed by normal background and then low voltage fast activity (LVFA), the UEO time would be marked at the beginning of the LVFA. For a stim seizure, if there is a clear “break” in the seizure during the stimulation and the time period before the break is not alone convincingly ictal itself, the expert was then instructed to call the beginning of the following sustained period the UEO (**Supplemental Material; Fig. S1**).

For the locations of seizure onset, the expert was instructed to list all channels showing an ictal pattern from the time of UEO to 1 second after (UEO + 1 second). For the locations of seizure spread after 10 seconds, the expert was instructed to list all channels showing an ictal pattern during the one second period after 10 seconds following UEO (from UEO + 10 seconds to UEO + 11 seconds). If a channel demonstrated an ictal pattern before the 10 second mark but did not occur during the 1 second window after 10 seconds, then the channel was not included in the spread annotation.

## DATA RECORDS

The dataset is hosted on <u>pennsieve.io</u>,^17^ a federated platform for data storage and analysis, and is publically available at (<link to open dataset upon publication>). The data closely follows the iEEG BIDS format,^24^ which contains a detailed description of the file structure. General metadata including BIDS version, licensing details, contributor information, and acknowledgments are documented in “dataset_description.json”. The dataset was generated in part using MNE BIDS.^25^ The data repository contains dataset-level metadata about patients in “participants.tsv” and about seizures in “annotations.tsv.” Each patient’s folder contains electrode localization derivatives as well as raw EEG EDF files (**Fig. 2**).

**Figure 2:**
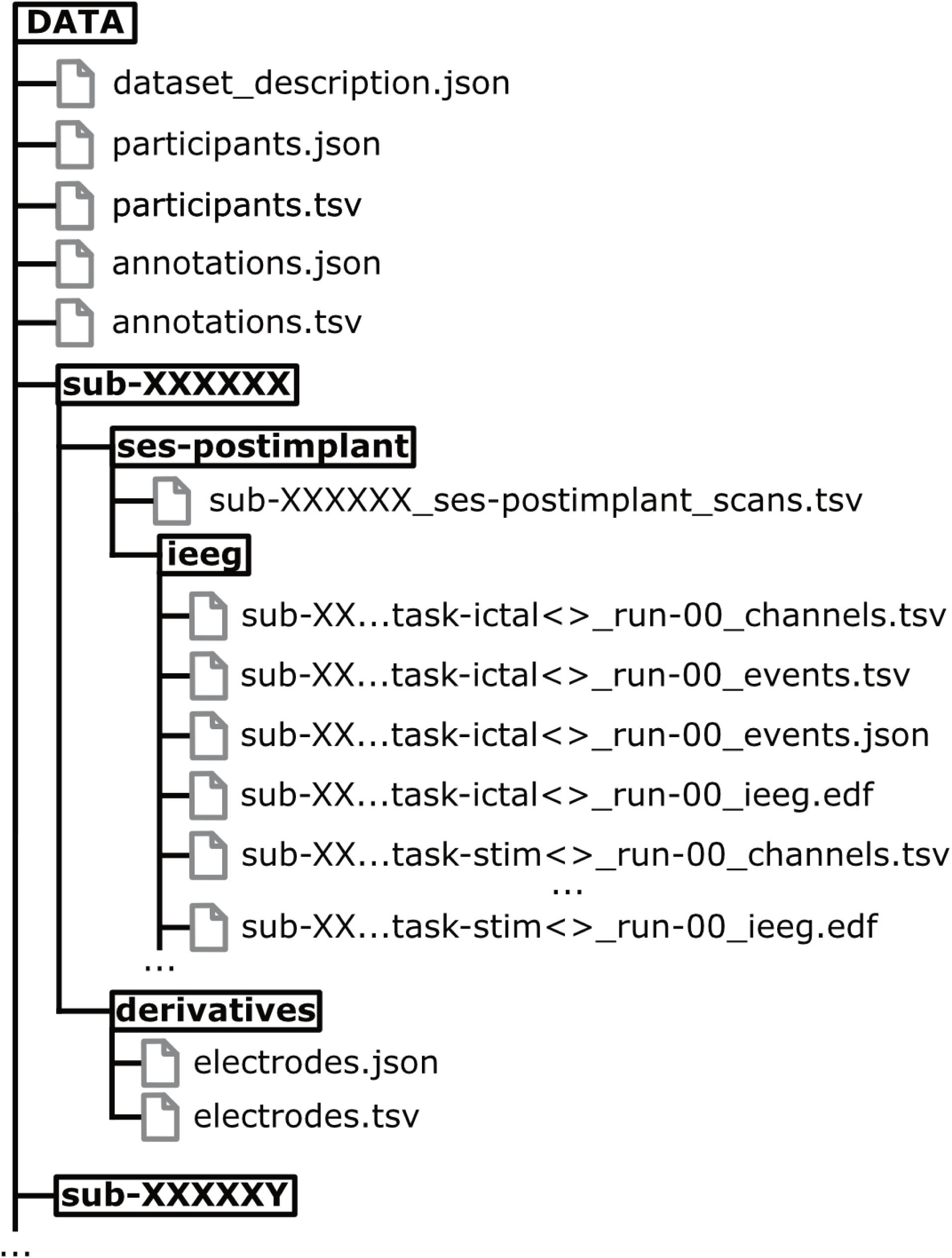
File hierarchy of the annotation repository.

### participants.tsv

This table contains participant-level metadata and is presented in **Table 1**. Each row represents a unique participant identified by “participant_id” (with prefix HUP indicating adult center and CHOP indicating pediatric center). Demographic variables include “age” at implantation (in years) and biological “sex” (F: female, M: male).

Clinical characterization variables describe the epilepsy phenotype and outcomes. The “mtle” field is a binary variable (Yes/No) indicating whether spontaneous seizures originated from mesial-temporal structures, consistent with mesial temporal lobe epilepsy (MTLE). The “unifocal” field describes whether the clinical impression based on recorded spontaneous seizures indicated a single focal epileptogenic zone (Yes) or a distributed network (No), where distributed networks include either diffuse seizure onset or multiple focal onset zones. For participants with non-unifocal onset zones, if mesial temporal structures were involved in the seizure onset zone, they were classified as having MTLE. The “lesional” field indicates whether epileptogenic abnormalities were detected on pre-surgical imaging (Yes: lesional, No: non-lesional).

Surgical outcome is described by the “outcome” field, which contains the International League Against Epilepsy (ILAE) score for post-surgical seizure freedom assessed at 24 months post-surgery (1: completely seizure-free; 2: only auras; 3: 1-3 seizure days per year; 4: 4 seizure days per year to 50% reduction). When outcome data were unavailable, this field is marked as “Unknown”.

Epilepsy history variables include “age_at_onset” (in years) and “duration” (in years), which together determine the age at implantation. Finally, “n_seizures” reports the total number of seizures included from that participant, of which “n_stim_induced” indicates the number induced by low-frequency electrical stimulation.

### annotations.tsv

This table contains seizure-level metadata describing both seizure characteristics and expert clinician annotations. Each row represents a single seizure event with annotations from three to five expert clinicians.

Seizure identification fields include “patient” (participant identifier indicating adult [HUP] or pediatric [CHOP] center) and “iEEG_ID” (the file name on the iEEG.org cloud storage platform).^26^ Temporal reference points are provided by “approximate_onset” and “end” (both in seconds relative to implant), which describe the reference times clinicians used for approximately when the seizure started and ended.

Seizure induction is characterized by the “stim” field, which indicates whether the seizure was induced by low-frequency electrical stimulation (“Stim. Induced”) or occurred spontaneously (“Spontaneous”). For stimulation-induced seizures, “stim_channels” specifies the electrode contacts where electrical stimulation was delivered. The “semiology” field provides additional classification for stimulation-induced seizures, indicating whether the induced seizure reproduced the participant’s habitual spontaneous seizure semiology (Typical), did not reproduce it (Atypical), or if classification was unavailable (Unknown). For spontaneous seizures, this field is marked as “Unknown”.

Expert annotation fields are structured to preserve both individual clinician assessments and consensus determinations. The “clinician” field contains a list of three to five entries identifying the annotating clinicians. For each seizure, clinicians independently marked the unequivocal electrographic onset (UEO) time and the channels exhibiting seizure activity at onset and at 10 seconds post-onset. Individual annotations are stored in list format: “ueo_time” contains each clinician’s marked onset time (in seconds), “ueo” contains the channels each clinician identified as seizing at onset, and “sec” contains the channels each clinician identified as seizing 10 seconds after their determined onset time.

Consensus annotations were derived from individual clinician assessments to establish reference standards while preserving information about inter-rater variability. The “ueo_time_consensus” field contains the consensus onset time calculated as the median of individual clinician annotations. The “ueo_consensus” and “sec_consensus” fields contain consensus channel sets determined by majority voting across individual clinician annotations. The consensus agreement reflects the gold-standard channel annotation, while the individual annotations preserve and quantify uncertainty in harder-to-annotate seizures.

### electrodes.tsv

This table is located in each patient’s derivatives folder and contains electrode localization data, with different localization methods used for pediatric (CHOP) and adult (HUP) participants reflecting differences in clinical workflows at each center. All tables include a “label” field containing standardized electrode contact labels for merging with raw iEEG data and clinical annotations.

CHOP electrode localizations provide CT space coordinates and anatomical parcellation. Spatial coordinates are given by “x”, “y”, and “z” fields in mm. The “matter” field specifies tissue localization of each electrode contact (grey: grey matter; white: white matter; CSF: cerebrospinal fluid; unknown: tissue type could not be determined). Anatomical localization is provided by the “brain_area” field containing Desikan-Killiany atlas regions of interest (ROI).

HUP electrode localizations provide comprehensive coordinate information across multiple spatial reference frames alongside standard FreeSurfer outputs. Spatial coordinates are provided in three coordinate systems: native space (“mm_x”, “mm_y”, “mm_z” in millimeters), FreeSurfer surface space (“surfmm_x”, “surfmm_y”, “surfmm_z” in millimeters), and voxel space (“vox_x”, “vox_y”, “vox_z” in voxels). This multi-space representation facilitates integration with various neuroimaging analysis pipelines. Anatomical localization follows the DK atlas, with “roi” specifying the DK region name and “roiNum” providing the corresponding numerical label from the FreeSurfer parcellation.

### iEEG

Each patient’s folder is marked by their subject ID (e.g., sub-HUP224), which can be used to link their raw EEG to their patient metadata. Each patient folder contains the session identifier, “ses-postimplant” and the data type folder “ieeg.” Within “ieeg,” seizures are organized as separate tasks, with the task as “ictal” for spontaneous seizures and “stim” for stimulation-induced ones. The task has the integer truncated approximate onset time concatenated after the seizure type (e.g. task-ictal202170) to link the raw EEG back to the seizure-level metadata in annotations.tsv. Each seizure contains sidecar events and channels files that contain the sampling frequency, channel names, and recording duration. To avoid introducing artifact or degrading information contained in the raw signal, we do not perform any channel rejection or signal preprocessing steps on the EEG data.

## TECHNICAL VALIDATION

To assess the quality of our clinician annotations and establish a gold-standard consensus, we quantify spatio-temporal annotation reliability using three complimentary approaches. First, to assess reliability of channel annotations, we calculated average pairwise inter-rater agreement by comparing each possible pair of annotators (interrater, **Fig. 3A**). Second, we calculated each individual annotator’s agreement with the consensus annotation (agreement with consensus, **Fig 3A**). Lastly, we estimated the temporal reliability of our clinician annotators, quantifying temporal consistency for each seizure **Fig. 4A**). All statistical comparisons were made with linear mixed-effects models to account for multiple seizures per patient, and, to reflect parametric testing, distributions are reported as a mean ± 95% CI. Because our annotators were experts in adult epilepsy, we report validation of expert annotations on patients from the adult epilepsy center in these primary analyses, though we provide seizures and annotations for all patients (**Supplementary Material**; **Fig. S3**).

**Figure 3:**
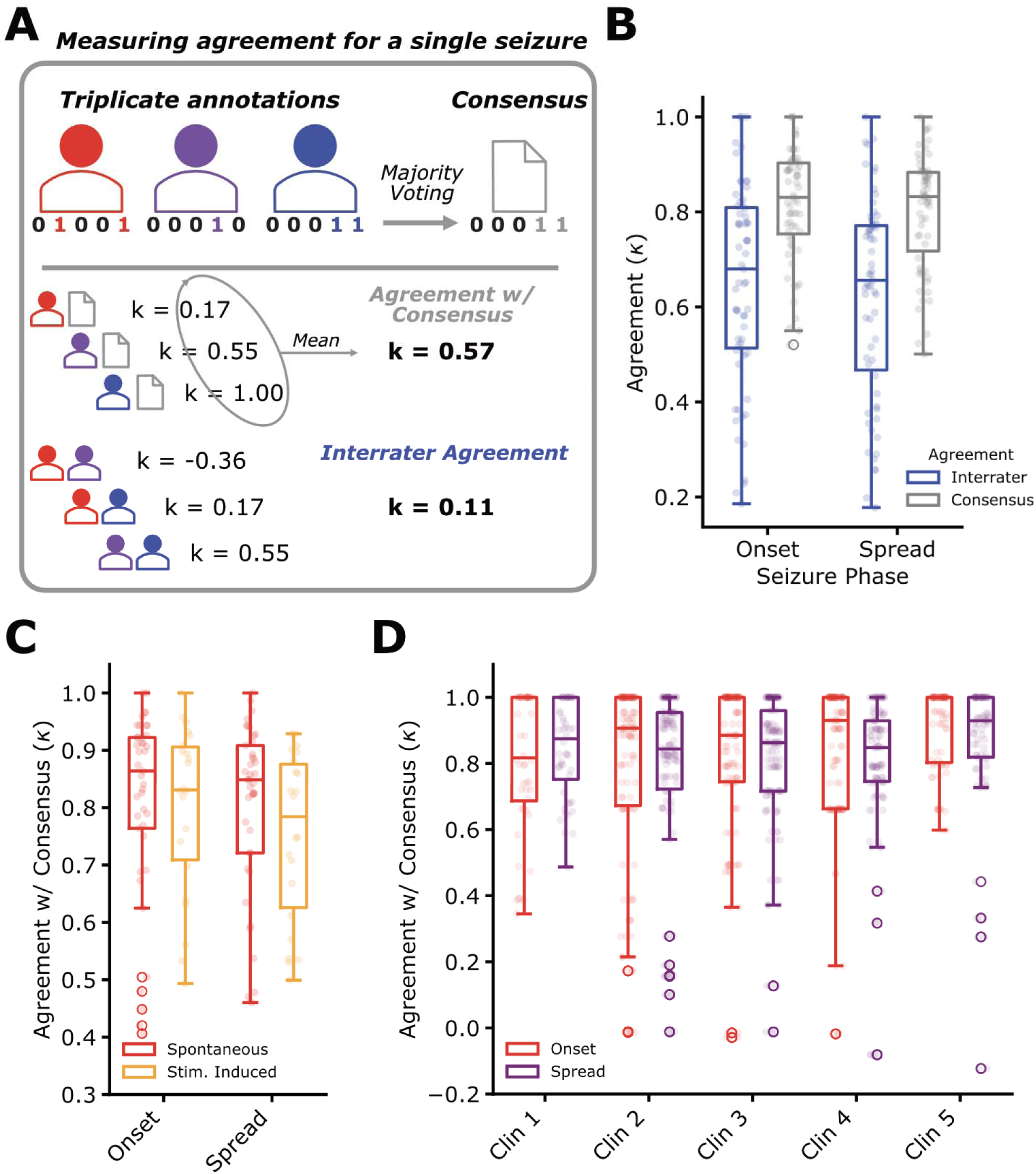
Seizure onset and spread channel annotator reliability. **A**) Example procedure for calculating EEG channel-level annotator agreement. For both onset and spread annotations on each seizure, we created a consensus annotation via majority voting. We then took the average Cohen’s kappa ( ) of each annotator with the consensus (Agreement w/ Consensus, gray) and between each annotator (Interrater Agreement, blue). **B**) For both onset and spread annotations, consensus was higher than interrater agreement (N = 63 seizures; Onset: = −0.17, SE = 0.027, z = −6.3, p = 3e-10; Spread: = −0.18, SE = 0.026, z = −6.8, p = 1.2e-11). **C**) Similarly, we saw consistent annotator agreement between stim (N = 21) and spontaneous (N = 42) seizures across both onset (β = −0.00, SE = 0.031, z = −0.15, p = 0.88) and spread (β = −0.03, SE = 0.031, z = −1.0, p = 0.30). **D**) To test whether annotator identity influenced consensus agreement, we fit mixed-effects models with patients as a random intercept. Across both onset and spread, the patient-level variance was extremely small (patient intercept variance ≈ 0.001). Given the negligible between-patient variance, we report the corresponding OLS results, which yielded nearly identical parameter estimates and revealed a nonsignificant effect (onset: F = 1.81, p = 0.13; spread: F = 1.11, p = 0.35), indicating that annotator identity does not meaningfully alter K values.

**Figure 4:**
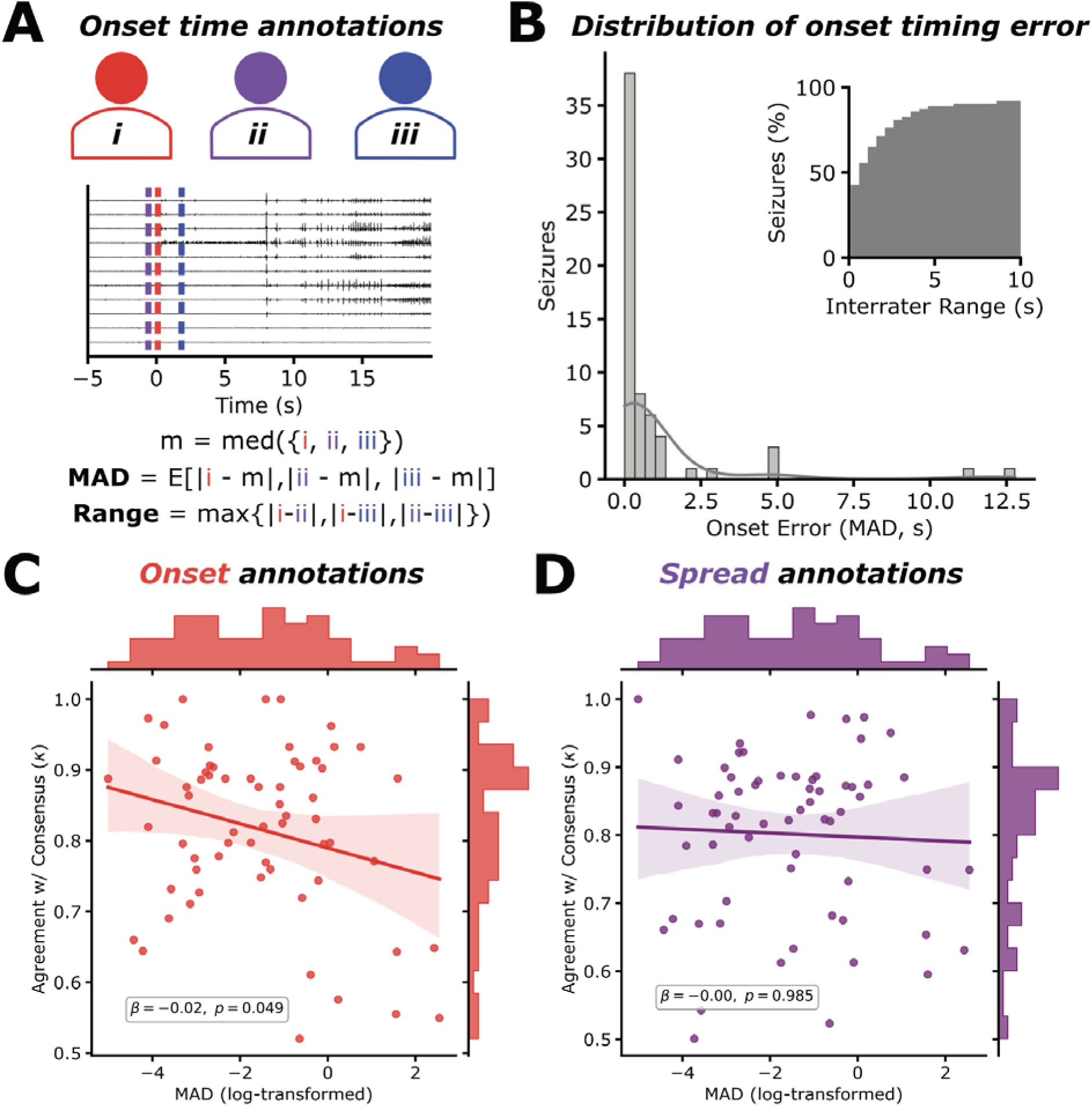
Seizure onset time annotation reliability. **A**) Schematic demonstrating example onset time annotations for three annotators, and the equations for calculating two measures of onset timing error: mean absolute deviation (MAD) and range. **B**) distributions of MAD and range (inset) demonstrating consistent onset timing annotations. **C**) Joint plot showing the relationship between onset timing error—log-transformed to normalize the distribution—and onset channel agreement. We observed a significant, negative correlation between the two features (= −0.017, SE = 0.01, z = −1.97, p = 0.049), though MAD only explained a small portion of the variance in onset agreement (marginal = 0.069; conditional = 0.11). **D**) We observed no significant relationship between onset timing and spread channel annotation agreement (= −0.00, SE = 0.01, z = −0.02, p = 0.985).

For channel selection (onset and spread), agreement was measured using Cohen’s kappa (κ), which accounts for chance agreement and ranges from −1 (perfect disagreement) to +1 (perfect agreement), with 0 indicating chance-level agreement. We saw that for both onset and spread annotations, interrater agreement (onset κ = 0.64 ± 0.06; spread κ = 0.62 ± 0.05) was significantly lower (p < 0.0001) than rater agreement with consensus (onset κ = 0.81 ± 0.06; spread κ = 0.80 ± 0.06) (**Fig. 3B**). This range is consistent with previously-reported levels of interrater agreement for seizure onset annotation,^8,9^ confirming “substantial” to “almost perfect” agreement between annotators,^27^ and provides a lower and upper bound on the expected agreement of expert human annotators. We saw no significant difference in reliability between stimulation-induced (onset κ = 0.81 ± 0.05; spread κ = 0.78 ± 0.05) and spontaneous seizures (onset κ = 0.82 ± 0.04; spread κ = 0.81 ± 0.04) (**Fig 3C**). Rater-consensus agreement did not differ across annotators (p > 0.10; **Fig 3D**), confirming that annotation reliability was stable across individuals.

To describe the relative consistency of onset time annotations, we calculated the mean absolute deviation (MAD) across the annotators’ marked times and the range (maximum temporal difference between any two annotators) (**Fig. 4A**). We saw that the majority of seizures had a total range of less than 1 second (53%), our margin for onset activity, and 87% of seizures were annotated within a total range of 5 seconds. Our annotations had a median MAD of 0.22 seconds, suggesting highly consistent temporal annotations. As expected, there was a significant relationship between onset time error (log-transformed MAD) and onset channel annotation agreement with consensus (p < 0.05; **Fig. 4C**). However, timing error only explained 7% of the variability in annotator onset channel agreement, implying that even when experts agree on the *timing* of seizure onset, they often disagree on the *channels* involved at onset reflecting the complex electrographic patterns occurring at seizure initiation. There was no significant association between spread channel annotation agreement and onset timing error (p = 0.99; **Fig. 4D**).

## DATA OVERVIEW

To illustrate evaluation methods enabled by multi-rater consensus annotations, we provide an example comparison of a single-channel seizure detection algorithm against clinician onset and spread annotations. The model was trained to detect seizure activity in one second increments on a single channel of intracranial EEG, and the signal preprocessing, WaveNet style architecture,^28^ and training protocol are described in previous work.^29^ We evaluate the model for its ability to discriminate between non-seizing and seizing channels at onset and spread and observe a strong AUC (onset: 0.83 ± 0.07; spread: 0.91 ± 0.05; **Fig 5A**) and relatively strong AUPRC (onset: 45 ± 0.11; spread: 0.52 ± 0.10; **Fig 5B**). However, when we compare predictions at a threshold optimized to detect seizure onset channels on an independent dataset, we see that the model matches the consensus annotation significantly worse than the interrater agreement on both onset and spread annotations (p < 0.0001; **Fig. 5C**).

**Figure 5:**
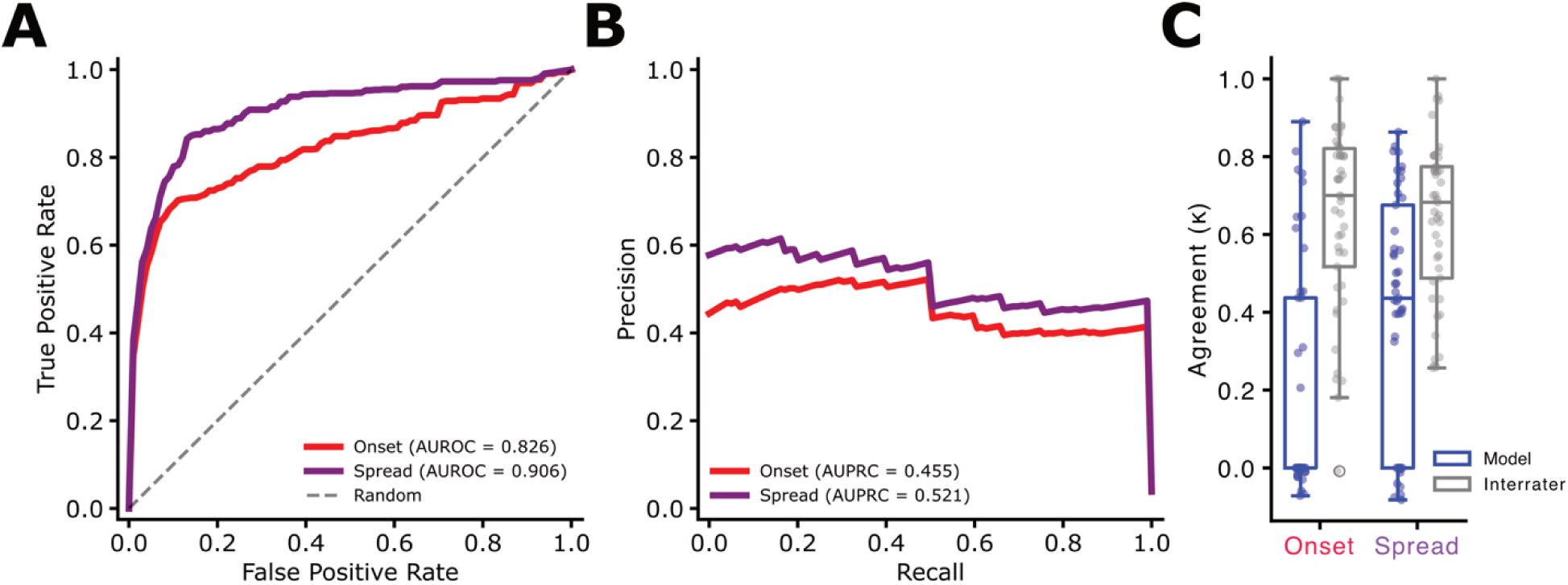
Benchmarking seizure annotation models. Here we show receiver-operator (**A**) and precision-recall (**B**) curves for annotating consensus seizing vs. non-seizing channels at the consensus seizure onset time (red) and after 10 seconds of seizure evolution (purple). **C**) Model agreement with clinician consensus was significantly lower than the interrater agreement in a paired comparison for both onset (β = −0.44, SE = 0.07, z = −6.76, p = 1.4e-11) and spread (β = −0.24, SE = 0.05, z = −4.66, p = 3.1e-6) annotations.

Here, we demonstrate a method for assessing model performance against our multi-annotator labels. The gold-standard consensus provides rigorous labels for a task even human experts can disagree on (**Fig. 3B**), and interrater reliability provides a positive control for when annotations are “good enough” to be considered human-level. Further, paired comparisons between model-consensus and interrater agreement account for variability in onset time annotations (which we show is related to interrater onset channel variability **Fig. 4C**), and the innate variability in clinical complexity across seizures and patients. This comparison highlights how the dataset enables standardized benchmarking of automated seizure annotation methods across both onset and spread detection tasks, as well as the outstanding work required to develop algorithms that reach the human-level agreement required to accelerate and implement clinical applications of automated methods.

## USAGE NOTES

The data can be downloaded from <u>pennsieve.io</u> and can be referenced here: <link to open dataset upon publication>. We also provide functionality to load the raw EEG data into a pandas dataframe from the BIDS configuration. Examples for how to preprocess the data can be found in our previous analyses using subsets of this data.^30^ EDF data can be streamed on Pennsieve, or clips can be downloaded partially or in full. Details on data download and API to interact with the data are previously described^17^ and on the Pennseive documentation hub (https://docs.pennsieve.io/docs/getting-started)

## Data Availability

The data can be downloaded from pennsieve.io and can be referenced here: (link to open dataset upon publication). All technical validation analyses and plots along with example code for loading the raw EEG from the BIDS dataset is publicly available https://github.com/wojemann/stim-seizures/tree/scientific_data. This repository also contains scripts used to generate the data overview analysis and subsequent figures, and the seizure annotation model along with an additional suite of pretrained or unsupervised seizure annotation algorithms is available as part of the DynaSD seizure annotation package (https://github.com/wojemann/DynaSD).

## CODE AVAILABILITY

All technical validation analyses and plots along with example code for loading the raw EEG from the BIDS dataset is publicly available https://github.com/wojemann/stim-seizures/tree/scientific_data. This repository also contains scripts used to generate the data overview analysis and subsequent figures, and the seizure annotation model along with an additional suite of pretrained or unsupervised seizure annotation algorithms is available as part of the DynaSD seizure annotation package (https://github.com/wojemann/DynaSD).

## Author contributions

**WKSO** and **DZ** wrote the original draft of the manuscript. **EC, BL, BCK, SKK, CA, EDM** performed data collection. **CVKC, DZ, EC, JK, JJL** acted as expert epileptologist annotators. **WKSO, EC, CA, NS** curated metadata and organized the open source dataset. **BL, EC** acquired funding for the study. **All authors** contributed to reviewing and editing of the final draft of the manuscript.

