## Supplemental Material for "Multi-expert consensus annotations of spontaneous and stimulation-induced seizures in stereotactic EEG"

Supplementary Material

### 1. Seizure annotation standard operating procedure

1. Overview

This protocol describes the standardized procedure used by three independent board-certified epileptologists to annotate seizure onset time and spatial localization in stereotactic EEG recordings. Annotations were performed for both spontaneous and stimulation-induced seizures to establish multi-expert consensus ground truth for algorithm development and validation.

2. Platform Access and Dataset Navigation

Annotations were performed using the iEEG.org cloud platform. Annotators created accounts and were granted access to de-identified patient datasets by the study coordinator.

For each seizure:

1. Open the dataset using the provided iEEG file identifier

2. Navigate to the approximate onset time provided in the metadata spreadsheet

3. Select an appropriate bipolar montage (clinical montages labeled “Montage 1”, etc., were typically used)

4. For stimulation-induced seizures, stimulating channels may be hidden for visualization purposes, but must still be considered for onset and spread annotations

3. Visualization and Filtering

Annotators reviewed recordings in bipolar longitudinal montage with all channels initially visible, with the option to hide selected channels as needed. Optional filtering was applied to facilitate interpretation when artifact obscured the signal.

3.1. Bandpass filtering for stimulation artifact:

For stimulation-induced seizures where DC drift and stimulation artifact rendered the EEG unreadable, annotators could apply a 3-50 Hz bandpass filter. In the iEEG.org interface, this was achieved by selecting Filters > Bandpass, setting low and high cutoff frequencies to 3 and 50 Hz, applying settings to all channels, and waiting several seconds for the filter to take effect.

3.2. Notch filtering:

For excessive 60 Hz artifact, annotators could apply a 59-61 Hz notch filter. Due to platform requirements, this required first applying a broad bandpass filter (0.5-256 Hz to all channels), then enabling the notch filter checkbox with cutoff frequencies of 59-61 Hz, and applying settings to all channels.

All filtering was applied uniformly across channels when used. The iEEG.org annotation layers could be toggled off (via the Layers button) if they obscured signal visualization. The viewer window timestamp (displayed in the “Start(s)” box when the window was aligned with the left edge of the screen) was used to determine precise timing for annotations.

4. Seizure Onset Definitions

4.1. Unequivocal electrographic onset (UEO)

The UEO represents the first time point at which there is a clear beginning to a continuous electrographic seizure / ictal pattern.

Annotators were instructed to:

1. Identify a period with unambiguous seizure activity

2. Move backward in time to identify the earliest point where ictal activity is certain and continuous with the later definitive seizure activity

3. Record the iEEG timestamp (in seconds) for this onset time

4.2. Seizure onset pattern characterization

After annotating seizure onset time and involved channels, annotators characterized the electrographic pattern using standardized descriptors.

Pattern classification:

A. Low voltage fast activity (LVFA):

·	Criteria: Frequency >13 Hz AND amplitude <10 µV

·	If criteria not met, use option (B) below

B. Frequency + pattern

·	Construct descriptor using: [Frequency term] + [Pattern term]

·	Frequency: delta (<4 Hz), theta (4-7 Hz), alpha (8-13 Hz), beta (13-30 Hz), gamma (>30 Hz)

·	Pattern terms: rhythmic activity, discharges

4.3. Exclusion criteria

Herald spikes or other electrographic changes that do not remain continuous with the subsequent seizure activity should not be marked as UEO. For example, if isolated spikes are followed by return to baseline before sustained low-voltage fast activity begins, the UEO should be marked at the onset of the sustained activity. For stimulation-induced seizures, if there is a clear temporal discontinuity (“break”) during stimulation, and the pre-break period is not convincingly ictal by itself, annotators were instructed to mark the beginning of the sustained post-break period as the UEO rather than earlier transient activity. Examples of appropriate and inappropriate labeling of UEO for a spontaneous seizure and stimulation-induced seizure are shown in Figures S1 and S2, respectively.

5. Spatial Annotations

5.1. Onset channels (UEO to UEO + 1 second)

Annotators identified all electrode contacts showing ictal activity during the 1-second window from UEO to UEO + 1 second. Channels were recorded using the format: lead, electrode number (e.g., LC1, LC2, LI1, LI2).

5.2. Spread channels (UEO + 10 to UEO + 11 seconds)

Annotators identified all electrode contacts showing ictal activity during the 1-second window from 10 to 11 seconds after UEO. Channels that began seizing but then stopped within the first 10 seconds were excluded from this annotation.

5.3. Volume conduction

Channels were considered less likely to represent true ictal activity if the electrographic activity lacked high-frequency or sharp components or was clearly time-locked and lagging behind ictal activity on other contacts that more clearly showed primary seizure involvement

### 2. Example onset annotations

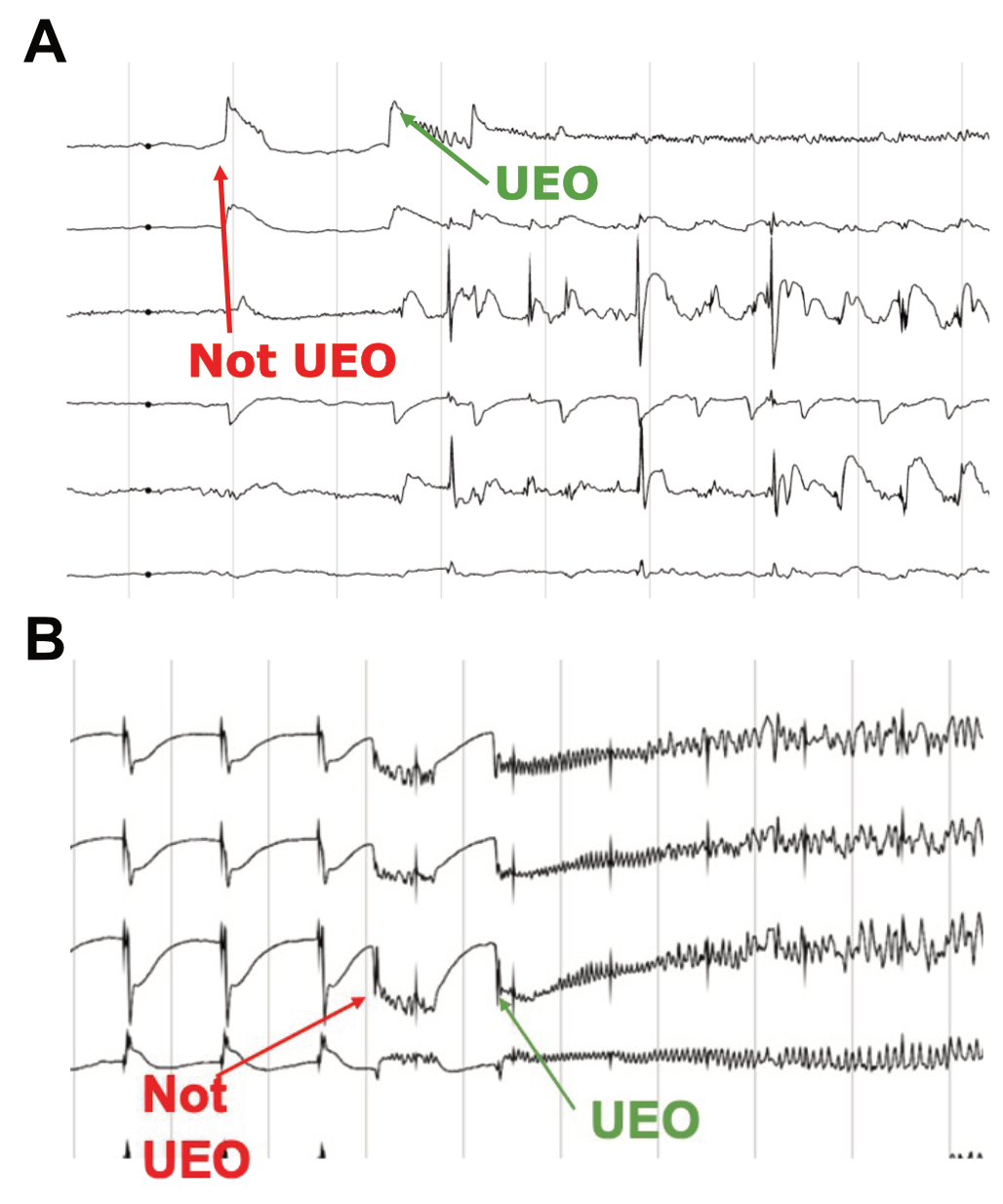

**Supplementary Fig. 1.** Example **A**) spontaneous and **B**) stimulation induced seizures demonstrating appropriate and inappropriate labeling of unequivocal electrographic onset (UEO) for this study.

### 3. Annotator agreement by center

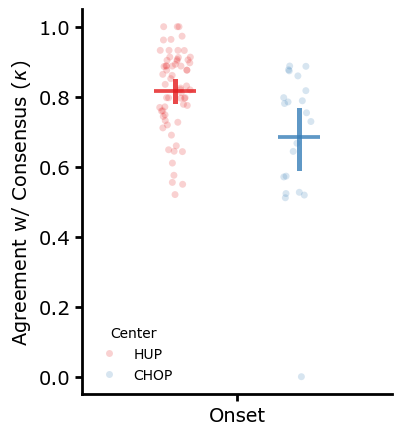

**Supplementary Fig. 2.** ***Center-level effects on rater–consensus agreement***

We compared agreement between individual annotators and the consensus across recording centers for onset labels. For onset annotations, a linear mixed-effects model with patient as a random intercept showed a significant effect of center (β = 0.131, SE = 0.039, z = 3.36, p = 0.001), with significantly higher agreement for HUP than CHOP. Mean κ values were 0.815 ± 0.021 for HUP and 0.684 ± 0.033 for CHOP. Based on this finding and the domain expertise of our annotators being adult epilepsy, we primarily focused our technical validation findings on solely the HUP patients.

### 4. Patient metadata table

| **participant_id** | **age** | **sex** | **mtle** | **unifocal** | **lesional** | **outcome** | **follow_up** | **duration** | **age_at_onset** | **n_seizures** | **n_stim_induced** |
| --- | --- | --- | --- | --- | --- | --- | --- | --- | --- | --- | --- |
| **HUP224** | 40-49 | F | Yes | Yes | No | Unknown | Unknown | 7.8 | 30-39 | 4 | 1 |
| **HUP225** | Unknown | M | No | Yes | No | 1.1 | 0.92 | Unknown | Unknown | 4 | 1 |
| **HUP229** | 40-49 | M | Yes | Yes | Yes | 1.1 | 3.92 | 32.9 | 0-9 | 2 | 1 |
| **HUP230** | 50-59 | F | Yes | Yes | Yes | 1.1 | 1.92 | 31.5 | 20-29 | 4 | 1 |
| **HUP235** | 20-29 | M | Yes | Yes | Yes | 1.2 | 1.5 | 10.9 | 10-19 | 4 | 1 |
| **HUP238** | 30-39 | F | Yes | Yes | No | 2.1 | 2 | 18.3 | 10-19 | 4 | 1 |
| **HUP246** | 50-59 | M | Yes | Yes | No | 1.1 | 2.08 | 4.3 | 40-49 | 2 | 1 |
| **HUP247** | 40-49 | F | No | Yes | Yes | 2.1 | 1.75 | 40.1 | 0-9 | 4 | 1 |
| **HUP249** | 40-49 | F | Yes | No | No | Unknown | Unknown | 2.8 | 40-49 | 4 | 1 |
| **HUP250** | 30-39 | F | No | No | No | Unknown | Unknown | 13.6 | 10-19 | 4 | 1 |
| **HUP253** | 40-49 | F | Yes | Yes | No | 1.2 | 1 | 17.2 | 20-29 | 4 | 1 |
| **HUP257** | 20-29 | M | Yes | Yes | No | Unknown | Unknown | 2.7 | 20-29 | 4 | 1 |
| **HUP261** | 20-29 | F | Yes | Yes | No | 1.1 | 1.17 | 28.9 | 0-9 | 2 | 1 |
| **HUP263** | 50-59 | F | Yes | Yes | No | 1.1 | 1 | 11 | 40-49 | 3 | 1 |
| **HUP266** | 30-39 | F | Yes | No | No | 2.1 | 0.75 | 9.3 | 20-29 | 4 | 1 |
| **HUP267** | 20-29 | M | Yes | No | No | Unknown | Unknown | 2.3 | 10-19 | 2 | 1 |
| **HUP273** | 30-39 | M | No | Yes | No | 2.2 | 1 | 12.2 | 20-29 | 3 | 2 |
| **HUP275** | 50-59 | M | No | No | No | 2.1 | 0.92 | 9 | 40-49 | 3 | 2 |
| **HUP288** | 30-39 | M | Yes | Yes | Yes | Unknown | Unknown | 16.1 | 10-19 | 2 | 1 |
| **CHOP005** | 0-9 | F | No | No | Yes | 1 | 4.54 | 1.3 | 0-9 | 1 | 1 |
| **CHOP010** | 10-19 | F | Yes | No | Yes | Unknown |  | 2.9 | 10-19 | 1 | 1 |
| **CHOP015** | 0-9 | M | No | No | Yes | 1.1 | 3.25 | 0.6 | 0-9 | 1 | 0 |
| **CHOP024** | 10-19 | F | No | Yes | Yes | Unknown |  | 13.1 | 0-9 | 1 | 1 |
| **CHOP026** | 10-19 | M | Yes | No | No | 1.1 | 2.17 | 2.8 | 10-19 | 2 | 2 |
| **CHOP028** | 10-19 | F | Yes | Yes | Yes | 2.3 | 2.01 | 1.8 | 10-19 | 1 | 1 |
| **CHOP035** | 10-19 | M | Yes | Yes | Yes | Unknown | Unknown | 2.4 | 10-19 | 2 | 2 |
| **CHOP037** | 20-29 | F | No | No | No | Unknown | Unknown | 11.4 | 10-19 | 2 | 2 |
| **CHOP038** | 10-19 | F | No | No | Yes | 3.1 | 1.14 | 12.9 | 0-9 | 2 | 1 |
| **CHOP041** | 10-19 | F | No | Yes | Yes | Unknown | Unknown | 7.1 | 0-9 | 2 | 1 |
| **CHOP045** | 10-19 | M | Yes | Yes | Yes | Unknown | Unknown | 0.6 | 10-19 | 1 | 1 |
| **CHOP046** | 10-19 | F | No | No | No | Unknown | Unknown | 8 | 10-19 | 2 | 2 |
| **CHOP049** | 0-9 | M | No | Yes | Yes | Unknown | Unknown | 1.2 | 0-9 | 2 | 1 |
